# Vagus Nerve and Stomach Synucleinopathy in Parkinson’s Disease, Incidental Lewy Body Disease and Normal Elderly Subjects: Evidence Against the “Body-First” Hypothesis

**DOI:** 10.1101/2020.09.29.20204248

**Authors:** Thomas G. Beach, Charles H. Adler, Lucia I. Sue, Holly A. Shill, Erika Driver-Dunckley, Shyamal H. Mehta, Anthony J. Intorcia, Michael J. Glass, Jessica E. Walker, Richard Arce, Courtney M. Nelson, Geidy E. Serrano

## Abstract

Braak and others have proposed that Lewy-type α-synucleinopathy (aSyn) in Parkinson’s disease (PD) may arise from an exogenous pathogen that passes across the gastric mucosa and then is retrogradely transported up the vagus nerve to the medulla. We tested this “body-first” hypothesis by immunohistochemically staining stomach and vagus nerve tissue from an autopsy series of 111 normal elderly subjects (no brain aSyn), 33 with incidental Lewy body disease (ILBD) (brain aSyn without clinical parkinsonism or dementia) and 53 with PD. Median disease duration for the PD group was 13 years. Vagus nerve samples were taken adjacent to the carotid artery in the neck. Stomach samples were taken from the gastric body, midway along the greater curvature. Formalin-fixed paraffin-embedded sections were immunohistochemically stained for α-synuclein phosphorylated at serine-129. In the vagus nerve none of the 111 normal subjects had aSyn in the vagus, while 12/26 ILBD (46%) and 32/36 PD (89%) subjects were aSyn-positive. In the stomach none of the 102 normal subjects had aSyn while 5/30 (17%) ILBD and 42/52 (81%) of PD subjects were aSyn-positive. As there was no aSyn in the vagus nerve or stomach of subjects without brain aSyn, these results support initiation of aSyn in the brain. The presence of aSyn in the vagus nerve and stomach of a subset of ILBD cases indicates that progression of synucleinopathy to the peripheral nervous system may occur at preclinical stages of Lewy body disease.

## INTRODUCTION

Since the discovery of α-synuclein (aSyn) as the major constituent of Lewy bodies, sensitive immunohistochemical (IHC) methods have demonstrated that the brain and peripheral nervous system (PNS) distribution and density of Lewy bodies and their associated abnormal neurites are much greater than formerly appreciated [1-4]. Furthermore, the common PNS occurrence of aSyn pathology has instigated a proliferation of research aimed at using PNS aSyn as a biopsy biomarker as well as raising the critical question as to whether or not aSyn begins in the brain or within the periphery [5-47]. The stimulus for the latter alternative, which might be called the “body-first” hypothesis, has come largely from clinical studies of PD that have found a wide range of non-motor signs and symptoms that accompany or may even precede the motor signs [48-50]. Many of these non-motor accompaniments are related to gastrointestinal (GI) dysfunction and therefore much attention has been focused on the stomach as the major “first stop” along the alimentary tract [43,51,52] and hence the most likely place for aSyn initiation, perhaps by absorption of toxins or through microbial or inflammatory stimuli, followed by transmission to the brain through the vagus nerve.

Supporting this is the repeatedly-confirmed finding of a rostrocaudal GI gradient of aSyn [1,12,14,31], which may itself be related to the known distribution of vagal GI innervation [53]. A stomach-vagal-brain connection has been invoked to explain reports that subjects with prior vagotomy may have a lower prevalence of PD, although this has been disputed [54-59]. A stomach “inoculation” site has now been experimentally tested in multiple animal models, and a consensus seems to have emerged that in fact there may be bidirectional spread both upwards from the stomach and downwards from the brainstem [60-62].

A major piece of evidence for the body-first hypothesis, however, has been lacking, in that human autopsy studies have never found pathology-specific forms of aSyn in the stomach or other GI location in the absence of such aSyn in the brain. Also, previous autopsy studies have focused on the GI tract itself but not on the supposed gut-to-brain conduit, the vagus nerve. It has been rightfully argued that since nervous tissue is much less concentrated in enteric walls than in CNS tissue, the apparent primacy of brain aSyn may only be due to its much greater endowment in this respect. The vagus nerve is, like CNS tissue, 100% nervous, and if it does serve as the connection through which aSyn makes its first passage from gut to brain, it, as well as a GI location, should be affected in a small percentage of normal subjects as the only manifestation of α-synuclein pathology. About 25% of clinically normal elderly subjects have a limited brain distribution of aSyn, most often in the olfactory bulb, brainstem and/or amygdala. These “incidental Lewy body disease” (ILBD) subjects also have reduced striatal dopaminergic markers [63-65], suggesting that they represent prodromal PD or dementia with Lewy bodies (DLB). If aSyn begins in the stomach and then passes through the vagus nerve to the brain, it might be expected that a similar percentage of normal older people would show aSyn limited to stomach and/or vagus nerve, but lacking aSyn in the CNS.

## MATERIALS AND METHODS

### Human subjects

The study took place at Banner Sun Health Research Institute (BSHRI), which is part of Banner Health, a non-profit healthcare provider centered in Phoenix, Arizona. BSHRI and the Mayo Clinic Arizona are the principal members of the Arizona Parkinson’s Disease Consortium. Brain necropsies and neuropathological examinations were performed on elderly subjects who had volunteered for the Arizona Study of Aging and Neurodegenerative Disorders (AZSAND)/BSHRI Brain and Body Donation Program (BBDP) [66]. The BBDP has been approved by designated BSHRI Institutional Review Boards and all subjects or their legally-authorized representative signed written informed consent. The majority of BBDP subjects are clinically characterized at BSHRI with annual standardized test batteries that include movement disorders and cognitive/neuropsychological components. Additionally, private medical records are requisitioned and reviewed for each subject and a postmortem questionnaire is conducted with subject contacts to help determine the presence or absence of dementia and parkinsonism for those subjects that did not have a recent standardized antemortem evaluation.

Subjects received complete neuropathological examinations while blinded to clinical diagnoses as described previously [66]. Specific consensus diagnostic criteria were used for PD, requiring 2 of 3 cardinal signs of rest tremor, rigidity or bradykinesia as well as substantia nigra aSyn and pigmented neuron loss at autopsy. Subjects with any brain aSyn but who lacked dementia or parkinsonism were termed incidental Lewy body disease (ILBD).

Case selection was done by searching the BBDP database for all those that had died and had a full clinical evaluation and full autopsy including vagus nerve and/or stomach sampling with immunohistochemical staining for phosphorylated α-synuclein. Vagus nerve samples were taken adjacent to the midpoint of the carotid artery in the neck while stomach samples were taken midway along the greater curvature.

### Histological methods

The process leading to the choice and evaluation of immunohistochemical methods for demonstrating pathological α-synuclein has been described in previous publications [16,67-70]. The standard method used at AZSAND employs proteinase K pretreatment, which not only results in superior epitope exposure but also may assist with the pathological specificity of the stain by digesting normal, non-aggregated α-synuclein, which is abundant in all nervous tissue. Using an antibody specific for α-synuclein phosphorylated at serine 129 (pSyn)[71-75] also helps identify stained structures as pathological since normal control subjects do not have pSyn-immunohistochemically positive brain tissue elements [4,76]. Complete details of the staining procedure have been previously described [77] and so only a brief description is given here.

From each postmortem subject, three sections from vagus nerve and three sections of stomach were stained and examined. Formalin-fixed, paraffin-embedded 5-7 μm sections were deparaffinized and treated with 1:100 proteinase K (Enzo Life Sciences, Farmingdale, NY) at 37**°** C for 20 minutes, followed by suppression of endogenous peroxidase activity with 1% hydrogen peroxide for 30 minutes, incubation in primary antibody against α-synuclein phosphorylated at serine 129, diluted 1:10,000 [72-75], incubation in biotinylated secondary antibody, avidin-biotin peroxidase complex (ABC, Vector Laboratories; Burlingame, CA) and 3,3’-diaminobenzidine (DAB; Sigma, St. Louis, MO) with saturated nickel ammonium sulfate and imidazole. All solutions subsequent to proteinase K, and all wash steps, excluding DAB incubation, were carried out in 0.1 M PBS with 0.3% Triton X-100, pH 7.4. Sections were then counterstained in 1% Neutral Red. Positive neuronal perikarya and nerve fibers are bluish-black while background and negative tissue structures are red. Cases were staged with the Unified Staging System for Lewy Body Disorders (USSLB) [3,4].

### Statistical tests

Group proportions in the three diagnostic groups were compared with chi-square tests. Group means were compared with one-way analysis of variance and post-hoc Newman-Keuls tests or two-way, unpaired t-tests.

## RESULTS

Subjects included 53 diagnosed clinicopathologically with PD, 33 with ILBD and 111 who were clinically non-demented without parkinsonism and had no CNS α-synuclein pathology (Table 1). The subjects were predominantly of advanced age, with the mean age ranges for the diagnostic groups falling between 83 and 87 years. Median disease duration for PD cases was 13 years, ranging from 2 to 44 years. PD cases were significantly younger (p < 0.05) than normal or ILBD cases, were more likely to be male when compared to the normal group (p = 0.006) and had higher UPDRS scores than the other groups (p < 0.0001). The mean postmortem intervals ranged between 4.1 and 4.6 hours and analysis of variance showed that there were no significant differences amongst groups. Vagus nerve was available for 26 ILBD, 36 PD and 111 normal control subjects. Stomach was available for 30 ILBD, 52 PD and 102 normal control subjects.

**Table 1.** Clinical characteristics of study subjects, by clinicopathological diagnosis, age, sex, last motor UPDRS score and disease duration. Means and standard deviations (SD) are given. ILBD = incidental Lewy body disease; PD = Parkinson’s disease; PMI = postmortem interval in hours; UPDRS = Unified Parkinson’s Disease Rating Scale, Part III; Dis Dur = disease duration in years. * PD cases were significantly younger (p < 0.05) than normal or ILBD cases, were more likely to be male when compared to the normal group (p < 0.05) and had higher UPDRS scores (p < 0.0001). The groups did not differ in terms of PMI.

| Diagnosis (N) | Age at Death* | Sex (% male)* | PMI | UPDRS* | Dis Dur |
| --- | --- | --- | --- | --- | --- |
| Normal (111)<sup>1</sup> | 85.1 (11.0) | 53.1 | 3.7 (2.5) | 7.6 (7.6)<sup>2</sup> | N/A |
| ILBD (33)<sup>1</sup> | 86.3 (8.2) | 57.6 | 4.5 (4.2) | 8.0 (6.2)<sup>3</sup> | N/A |
| PD (53)<sup>1</sup> | 79.1 (6.4) | 75.5 | 4.1 (2.6) | 38.8 (20.5)<sup>4</sup> | 13.3 (7.3) |

1. Not all subjects had both vagus nerve and stomach available. See text for details. 2. For 15 normal cases, UPDRS scores were not available. 3. For 5 ILBD cases, UPDRS scores were not available. 4. For PD, 16 scores were done while on medication, 35 while off medication; 2 were not done.

As in our previous investigations [1,17,77-79], only staining that was morphologically consistent with nerve fibers was considered to be specific for stomach or vagus nerve aSyn. Immunoreactive nerve fibers were present within the stomach or vagus nerve of 42/52 (81%) and 32/36 (89%) PD subjects, as compared to 5/30 (17%) and 12/26 (46%) of ILBD subjects (Table 2). No aSyn was present in stomach (102 subjects) or vagus nerve (111 subjects) of control subjects. Immunoreactive nerve fibers, when present in PD and ILBD subjects, were mostly sparse but occasionally focally frequent in the vagus nerve, and sometimes had abnormally large swollen segments, consistent with dystrophic change (Figure 1a, b). In the stomach, immunoreactive nerve fibers were found in all layers (Figure 2c-f) but most frequently were found in the submucosa, where they were often closely applied to the external surface of arterioles or within small nerve fascicles.

**Table 2.** Neuropathological characteristics of Lewy body disease study subjects, by vagus and stomach aSyn status. Means and standard deviations (SD) are given. ILBD = incidental Lewy body disease; PD = Parkinson’s disease.

| Diagnosis | USSLB Stage Mean (SD) | Brain aSyn Load Mean (SD) |
| --- | --- | --- |
| ILBD: Vagus aSyn + (n=12) | 2.33 (0.49) | 12.08 (8.57) |
| ILBD: Vagus aSyn - (n=14)<sup>1</sup> | 1.92 (0.64) | 6.08 (6.79) |
| ILBD: Stomach aSyn + (n=5) | 2.6 (0.55) | 12.2 (3.96) |
| ILBD: Stomach aSyn - (n=25)<sup>1</sup> | 1.96 (0.69) | 7.92 (8.30) |
| PD: Vagus aSyn + (n=32)<sup>1</sup> | 3.44 (0.67) | 26.93 (6.28) |
| PD: Vagus aSyn - (n=4) | 3.25 (0.96) | 25.75 (11.56) |
| PD: Stomach aSyn + (n=42)<sup>2</sup> | 3.62 (0.54) | 29.84 (5.83) |
| PD: Stomach aSyn - (n=10) | 3.0 (0.67) | 22.33 (5.81) |

1. Missing USSLB stage and brain aSyn load for one case. 2. Missing brain aSyn load in three cases.

**Figure 1.**
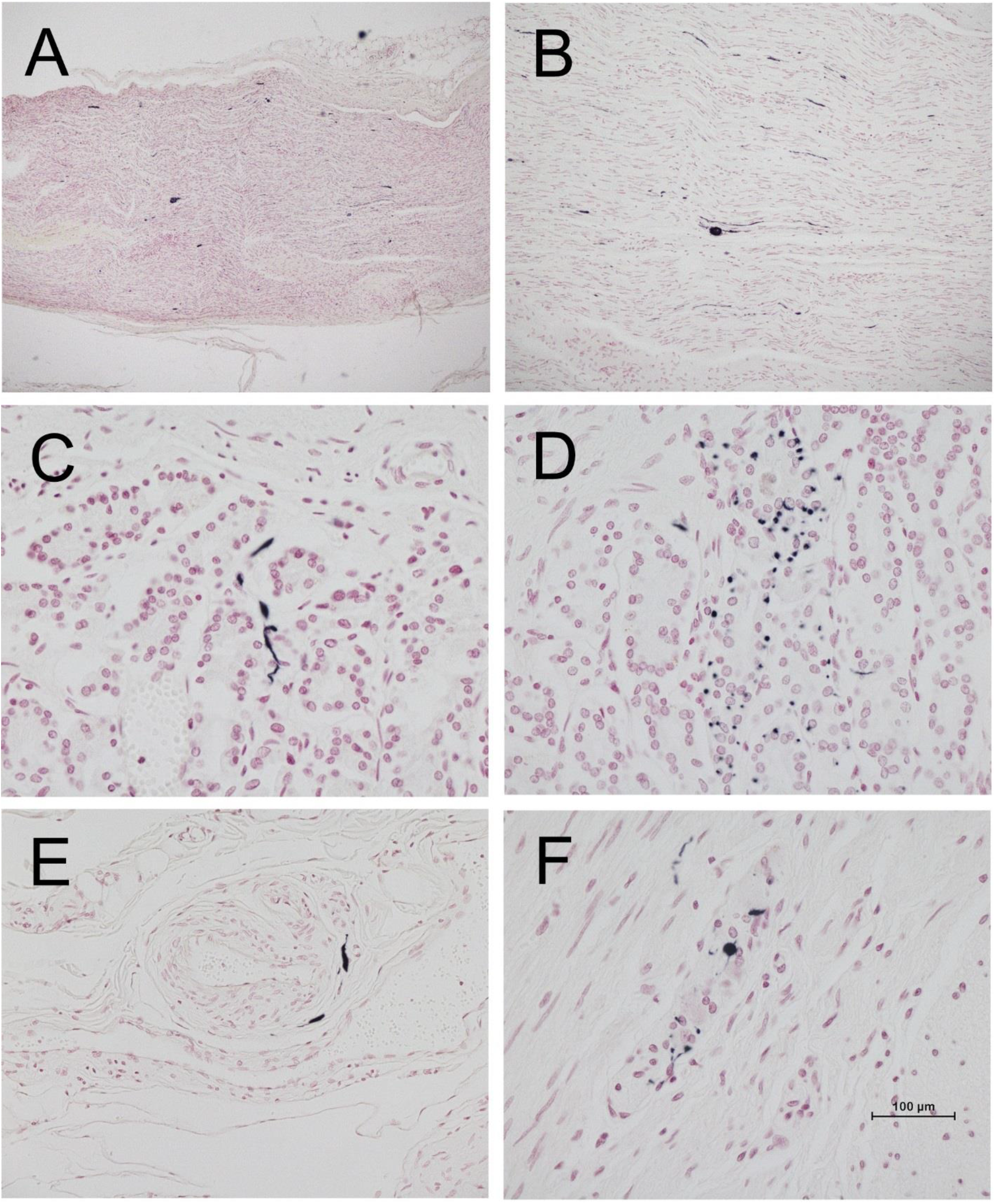
Photomicrographs of vagus nerve and stomach immunohistochemically stained for phosphorylated α-synuclein (black) and counterstained with Neutral Red (red). A and B are longitudinal sections of vagus nerve at low (A) and medium (B) magnification. C and D show short fibers and puncta in the stomach mucosa. E shows short fibers applied to the peripheral surface on an arteriole in the submucosa. F shows puncta within an intermyenteric ganglion. Calibration bar in F represents 100 μm for C-F, 400 μm for B and 800 μm for A.

Cases, both ILBD and PD, that were aSyn-positive in stomach or vagus tended to have higher brain Unified LB stages and higher total brain aSyn loads than those that were aSyn negative, and, on average, ILBD cases with vagus or stomach aSyn were in USSLB Stage IIa or IIb (brainstem or limbic-predominant) while PD cases were most often in Stage III (brainstem and limbic), although some ILBD cases were in Stage III and some PD cases were in Stage IV (Table 2). All cases with vagus or stomach aSyn were in Unified Stage IIa (brainstem predominant), IIb (limbic predominant) or higher.

## DISCUSSION

This is the first comprehensive assessment of the prevalence of stomach and vagus nerve aSyn in PD, ILBD and control subjects. The results show that stomach aSyn was present in 81% and vagus nerve aSyn was present in 84% of autopsy confirmed PD subjects, while in ILBD, aSyn was present in stomach in only 17% and in vagus in only 46% of subjects. There was no aSyn found in either the stomach or vagus nerve from any of the more than 100 clinically normal control subjects without brain aSyn. The lack of aSyn in stomach and vagus nerve, in subjects without any brain aSyn, as well as the lesser prevalences of aSyn in stomach or vagus nerve as compared to brain (all of the PD and ILBD cases had brain aSyn), suggest that the spread of aSyn to the stomach and PNS occurs subsequent to the establishment of threshold brain aSyn loads. USSLB Stages and total brain aSyn loads were higher in ILBD and PD cases with stomach or vagus aSyn.

A critical question has been whether or not aSyn begins in the brain or within elements of the PNS [80-82]. The present results do not support the concept that the initiation of α-synuclein pathology in Lewy body disorders begins in the PNS rather than the brain. Non-motor accompaniments of PD may predate or occur early in the motor progression [49,83-86] but the results for ILBD subjects suggest that even in these clinically prodromal subjects, aSyn has already been established in the brain. Autopsy studies of relatively small numbers of subjects with ILBD have demonstrated a high prevalence of aSyn within the spinal cord, sympathetic ganglia, adrenal medulla and upper GI tract [1,7,11,26,42,43] but no study to date has found aSyn in the spinal cord or in PNS sites in the absence of brain involvement, with the possible exception of Fumimura et al (2007) [45] who reported one case out of 783 with adrenal medulla as the only site with aSyn. One other case report, by Miki et al (2009) [87] exists of Lewy body pathology restricted to the heart and stellate ganglion.

Although we used an immunohistochemical method that has been repeatedly demonstrated to be highly sensitive and specific for both CNS and PNS α-synuclein pathology, as found in multi-center blinded studies [69,70,78,88] and biochemical studies [75], it is possible that the initial form of peripheral α-synuclein pathology may differ from that commonly seen in the CNS. Alternate forms may include non-phosphorylated α-synuclein, truncated α-synuclein [89-91] and α-synuclein aggregates [39, 92]. Also, conversion of normal α-synuclein to pathological varieties may occur more commonly in PNS locations other than stomach, perhaps on the basis of locally high normal α-synuclein concentrations [19], or due to inflammation [93-95]. It is possible that the initial “seeding” of aSyn, whether from the environment or by spontaneous internal generation, is accomplished by transiently-existing forms that may not locally incite the more familiar forms of aSyn but do so in the CNS once transmitted there. Assessment of PNS with α-synuclein seeding assays such as RT-QuIC or protein misfolding cyclic amplification (PMCA) may be more sensitive than IHC and may be more effective at uncovering PNS aSyn [96, 97]. Recent studies have suggested that bidirectional spread of α-synuclein aggregates may not necessarily pass through the vagus nerve but rather via a hematogenous route [61].

These results are most consistent with a first appearance of α-synuclein pathology in the CNS, with later spread, often at a premotor stage, to the PNS. All cases with either vagus or stomach aSyn were in Unified Stage II (IIa or IIb) or a higher stage.

## Data Availability

All data referred to in the manuscript is available on request from the corresponding author.

## ACKNOWLEDGMENTS

This study was funded by grants from the National Institute of Neurological Disorders and Stroke (U24 NS072026) the National Institute on Aging (P30 AG19610), the Arizona Department of Health Services (contract 211002), the Arizona Biomedical Research Commission (contracts 4001, 0011, 05-901 and 1001) and the Michael J. Fox Foundation for Parkinson’s Research.

## Notes

### Competing Interest Statement

The authors have declared no competing interest.

### Funding Statement

National Institute of Neurological Disorders and Stroke U24 NS072026. National Institute on Aging P30 AG19610. Arizona Department of Health Services. Arizona Biomedical Research Commission. Michael J Fox Foundation.

### Author Declarations

Western Institutional Review Board, Puyallup, WA

### Summary of Updates

The institutional affiliation for Charles Adler was incorrect in the original version. He is affiliated with the Mayo Clinic, not Barrow Neurological Institute. His affiliation has been changed to Mayo Clinic in the revised version. This necessitated a re-coding of several of the other authors but all the other authors had correct affiliations in the original version.

